# COVID-19 mortality: positive correlation with cloudiness but no correlation with sunlight and latitude in Europe

**DOI:** 10.1101/2021.01.27.21250658

**Authors:** S. Omer, A. Iftime, V. Burcea

## Abstract

We systematically investigated an ongoing debate about the possible correlation between SARS-CoV-2 (COVID-19) epidemiological outcomes and solar exposure in European countries, in the period of March – December 2020. For each country, we correlated its mortality data with: i) objective sky cloudiness (as cloud fraction) derived from satellite weather data, ii) solar insolation (watt/square metre at ground level), iii) latitude. We found a positive correlation between the monthly mortality rate and the overall cloudiness in that month (Pearson’s r(35)=.68, P<.001; averaged linear model fitting the data, adjusted R2 =0.45). In an additional month-by-month analysis, 17% to 59% of the variance in COVID-19 mortality/million appears to be predicted by the cloudiness fraction of the sky, except in the last two months of 2020. We did not find any statistical significant correlation with insolation, nor with latitude of the countries, when the “latitude of a country” was precisely defined as the average landmass location (country centroid). The unexpected correlation found between cloudiness and mortality could perhaps be explained by the following: 1) heavy cloudiness is linked with colder outdoor surfaces, which might aid virus survival; 2) reduced evaporation rate; 3) moderate pollution may be linked to both cloudiness and mortality; and 4) large-scale behavioural changes due to cloudiness (which perhaps drives people to spend more time indoors and thus facilitates indoor contamination).

## Introduction

In the initial year of the SARS-CoV-2 (COVID-19) pandemic, when effective countermeasures were not yet available (vaccination, antiviral therapy), the environmental factors that influenced viral transmission and mortality were hotly debated. This study explores the influence of previously ignored climatological factors (atmospheric cloudiness and solar irradiation) on COVID-19 mortality in Europe.

The factors that might influence the spread and impact of COVID-19 have been extensively discussed in the media and in research articles. Coronaviruses spread in the population due to a combination of medical, biological and socio-economic factors. It has also been suggested that latitude (1,2) and climate (temperature, humidity (3,4)) significantly contribute to COVID-19 spread.

A copious amount of (mainly local) studies (some not peer-reviewed) on climatic influence on COVID-19 has been released, with mixed or contradictory results. The World Meteorological Organization even hosted an international virtual symposium on the issue to deal with these uncertainties (“Climatological, Meteorological and Environmental factors on COVID-19 pandemic”, 4-6 August 2020).

Initially, (March – April 2020), the transmission of COVID-19 seemed to be associated with the 30- to 50-degree North longitude corridor and weather patterns and low specific and absolute humidity (2); however this could just as plausibly reflect trade and human movement patterns in the Northern hemisphere. Late in the spring of 2020, tropical and subtropical countries began to see an increase of pandemic transmission, and the latitude dependency thus appears to be unsure at this point.

It has also been previously noted that COVID-19 had a higher impact in countries where epidemiological data had shown a degree of vitamin D deficiency in the population (1). Vitamin D has a wide range of immunomodulatory, anti-inflammatory and antioxidant properties, and it thus seems (presumably) protective. In vivo vitamin D synthesis is photochemically dependent in humans, and its concentration levels drop without sufficient sunlight exposure. As the latitude increases, the amount of locally received sunlight generally decreases. These two facts were combined by other researchers in a range of compelling clinical hypotheses linking susceptibility to COVID-19 to vitamin D and indirectly to lack of sunlight exposure or increased latitude (see for example (5) or (6) for a more thorough perspective on these issues and (7) for a biochemical study).

In sharp contrast to the above hypotheses, other biochemical studies have not to found a correlation between vitamin D and COVID-19 epidemiological data (see (8) or (9)). These studies concluded that other unknown factors might be at play.

We tried to address these contradictions from an analytical, biophysical point of view: the amount of UV radiation that reaches the ground (and is thus presumably protective) is a fraction of total sunlight (which also contains visible light and infrared radiation). The total sunlight (known also as “solar insolation” or “solar irradiance at ground level”) is defined as the flux of solar radiation per unit of horizontal area for a given location. Solar irradiance is expressed in watts/square metre. It depends on several factors: primarily on solar zenith angle (which depends on latitude), secondarily on atmospheric composition (via absorption and scattering), and thirdly on seasonal change (due to the Earth’s axial tilt). These are well known and researched topics; see for example (10) for an extensive review. Solar irradiance is one of the main factors that determine the temperature at ground level (with an almost linear dependence (11)), along with humidity (by inducing water evaporation and soil dryness) and the climate in general (12)).

Solar irradiance at ground level is heavily influenced by atmospheric composition, primarily by clouds (13) and secondarily by other factors such as dust, pollutants, and humidity (14). Clouds exert a complex influence: by reflection of the sunlight (back into space) they reduce the amount of total energy that reaches the earth but by scattering they can counter-intuitively direct some of the energy back to land, especially in the UV portion of the spectrum, thus modulating the biologically effective radiation dose received by living things (15).

In this study, we present our detailed research on the direct influence on COVID-19 mortality in European countries of the above-discussed factors: a) sky cloudiness, b) solar irradiance and c) latitude. Our results show that among these three factors, there is a sizeable influence due to cloudiness and basically no observable influence due to the absolute amount of sunlight nor to latitude.

## Methods

### 1. Datasets

#### Epidemiological data

Different epidemiological variables about COVID-19 epidemics are collected and reported around the world. We used the “Coronavirus update”, a publicly available epidemiological database summarized by Worldometers (16); monthly data snapshots of it were retrieved from the Internet Archive Organization (17). From this database we extracted the mortality, defined as the number of deaths per 1 million inhabitants in a given country in a particular month. There are two issues with this approach that might impact our study. First, there are legal and practical differences among countries regarding death recording and reporting (18). The COVID-19 database we used records the deaths as reported by local authorities; we could not quantitatively assess the differences between the countries, which could impact comparison of different countries. Second, the time lag between the actual death and the reported time could be different for each case; the monthly intervals analysed probably average most of the differences.

#### Atmospheric cloudiness data

Cloudiness (also known as cloud fraction, cloud cover, cloud amount or sky cover) refers to the fraction of the sky obscured by clouds (in a particular location). It can be reported in various units. In this study, we used the cloud fraction (as tenths of the entire sky); 0.0 thus indicates a clear sky and 1.0 (or 10/10) indicates a completely covered sky.

We used a publicly available dataset of global cloudiness as measured from space by NASA’s Terra and Aqua satellites using the MODIS instrument (Moderate Resolution Imaging Spectroradiometer) (19). This dataset is collected continuously and presented as values averaged daily, weekly and monthly for the entire globe; for this study we chose the monthly averaged values.

In this dataset the entire Earth surface is divided into a rectangular grid. Each rectangle of the grid contains the average cloud fraction of the sky covering that area. The data are available at different resolutions (sizes of the rectangular grid that covers the Earth). We chose a fairly detailed resolution of the grid, 0.25° latitude x 0.25° longitude. We ensured that the other geographical datasets that we used in this study matched the same spatial resolution.

#### Solar data

The average solar insolation (also known as solar irradiance, solar exposure, incoming sunlight) in watts/square metre at the Earth’s surface was used in this study. We used a publicly available dataset inferred from measurements taken by Clouds and Earth’s Radiant Energy System (CERES) instrument flying aboard NASA’s Terra and Aqua satellites (20). We used the same temporal and spatial sampling as presented above, i.e., monthly averaged values over a 0.25° latitude x 0.25° longitude grid.

#### Geographical data

For a single point on the Earth, the geographical coordinates are straightforward (latitude and longitude). However, for a country (or large region) the coordinates can be represented in several distinct ways, each with pros and cons, such as: the location of the capital city, the location of the most populated city, the average between the most extreme points of the country, and the country centroid, among others.

As the “latitude of a country” measure, we used the country centroid. A centroid (also known as the centre of gravity or centre of mass) is the arithmetic mean of the positions of all the points in a geometrical object; for irregular objects, it is closest to the centre of the biggest part of the object (it is less influenced by very thin or heavily scattered boundaries). We chose this measure because several countries have highly irregular geometrical boundaries or long thin peninsulas or numerous islands (i.e., Greece, Norway, etc.); the country centroid is located closer to the widest area of the mainland. We used a standard database of countries centroids published by Google Maps developers (21).

As the “country boundary” measure we chose to use a simplified representation of the country border, the country bounding box, which is a rectangle on the surface of the Earth with North and South edges corresponding to the limiting (max and min) latitudes of the country and West and East edges corresponding to the limiting longitudes. We used a database of bounding boxes of all countries published by the Center of Humanitarian Data (22).

We are aware that the use of the country bounding box simplification can induce a degree of imprecision (smoothing) or overlapping errors. To minimize these errors, we carefully curated the bounding box coordinates to exclude non-mainland portions. We did this because we wanted to restrict the analysis to the land portion of Europe (for example, overseas remote islands such as Svalbard would include almost 600 sq km of Arctic Ocean surface in Norway’s geographical definition). We checked this by comparing the centroid location of each country with the average geometric centre of its bounding box, making sure that the error was less than 1 degree of longitude or latitude.

#### Inclusion and exclusion criteria

The inclusion criteria were as follows: a) all countries on the European continent; b) availability of epidemiological and geographical (cloudiness, insolation) data.

The exclusion criteria were as follows: a) population<0.5 million and b) a geographical bounding box of the country smaller than 0.25° latitude x 0.25° longitude. We used these criteria because European micro-states (Monaco, Vatican, San Marino, etc.) were too small to be properly sampled from the available resolution of geospatial data (insolation, cloudiness).

Russia was also excluded from analysis because the COVID-19 epidemiological data from the country was publicly available only in an aggregate form (i.e., no data were available detailing the epidemiology in European and Asian parts of Russia).

In this way we obtained a list of 37 European countries (listed alphabetically): Albania, Austria, Belarus, Belgium, Bosnia and Herzegovina, Bulgaria, Croatia, Czech Republic, Denmark, Estonia, Finland, France, Germany, Greece, Hungary, Ireland, Italy, Latvia, Lithuania, Luxembourg, Malta, Moldova, Montenegro, Netherlands, North Macedonia, Norway, Poland, Portugal, Romania, Serbia, Slovakia, Slovenia, Spain, Sweden, Switzerland, United Kingdom (UK) and Ukraine.

For each of these 37 countries and for each one of the 10 months in the studied interval (March – December 2020), we included in the analysis the following variables: i) COVID-19 mortality (deaths per 1 million population), ii) insolation averaged for 1 month across the country (measured in Watt/m2), iii) the cloud fraction averaged for 1 month across the country (expressed as a fraction between 0.0 … 1.0) and iv) the latitude of the countries (centroid data). Thus the total number of aggregated datapoints was 1480; we made this database freely available (see Data Availability section). As a convenience for the reader we included a quick visual overview of these aggregated results the Supplementary materials, Figures S.1, S.2 and S.3)

### 2. Software used

We performed basic epidemiological data gathering in tabular sheets (*Excel/LibreOffice Calc*). We performed data analysis and visualization in *R* version 4.0.2 (23), with packages *tidyverse* (24), *ggplot* (25), *stargazer* (26), *lmtest* (27); geospatial data was analysed with *raster* package (28).

### 3. Calculation

Most of the European countries reported the first deaths in March 2020 (see Supplementary material, Figure S.1), and we used this month as the starting point for our analysis.

We employed multiple linear regression models with mortality as the response (it is log10 transformed in all the following analyses and figures). As predictor variables we used cloud fraction and solar insolation, both averaged (for the March – December interval). For each one of the reported models we checked their validity by analysing the normality of the residuals as follows: normal Q-Q plot, presence of outliers, and homogeneity of the variance of residuals (we assumed homoscedasticity if the result of the studentized Breusch-Pagan test was bigger than 0.05). We attempted a brief time-series analysis of the factors; we checked the distribution of the data in time and the autocorrelation of the disturbances with a Durbin-Watson test.

## Results

### 1. Overall Mortality vs. Cloudiness

For each country, in the interval March – December 2020, we averaged the monthly cloudiness and the monthly mortality rate, and we built a linear model (Figure 1). It shows a modest but statistically significant correlation between the average mortality rate and the average sky cloudiness (Pearson’s r(35)=.68, P<0.001).

**Figure 1.**
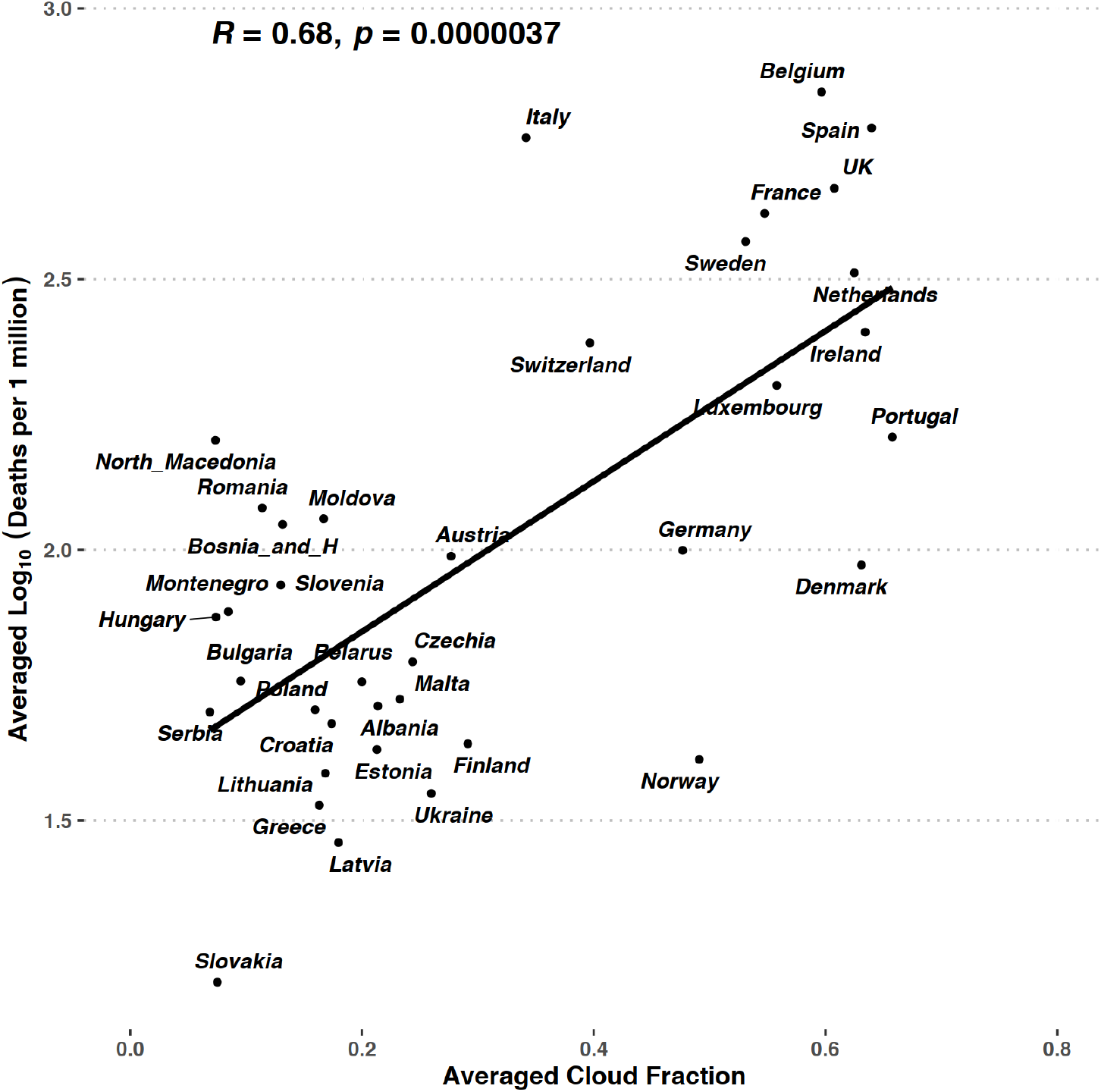
Overall correlation between mortality and cloudiness. The data points are the averaged values for each one of the European countries included in the study during March – December 2020. Cloudiness is expressed as the cloud fraction, ranging between 0.0 (completely sunny sky) and 1.0 (clouds completely obscure the sun).

The linear model that fits the data shown in Figure one is presented in Table 1. In continental Europe, from the beginning of the epidemic, approximately 45% of the variance in COVID-19 mortality/million appears to be predicted by the cloudiness fraction of the sky. The model passes the quality criteria listed in the Calculation section, and the residuals of the model appear to be homoscedastic, Breusch-Pagan test P-value=0.492).

**Table 1.** The statistics of the linear model of the overall averaged mortality vs. averaged cloudiness.

| *Dependent variable:* |  |
| --- | --- |
| Averaged Log<sub>10</sub>(Deaths/million) |  |
| Averaged Cloud_Fraction | 1.389*** |
|  | (0.253) |
|  | t = 5.488 |
|  | p = 0.00001 |
| Constant | 1.571*** |
|  | (0.094) |
|  | t = 16.691 |
|  | p = 0.000 |
| Observations | 37 |
| R<sup>2</sup> | 0.463 |
| Adjusted R<sup>2</sup> | 0.447 |
| Residual Std. Error | 0.313 (df = 35) |
| F Statistic | 30.118*** (df = 1; 35) (p = 0.00001) |
| *Note:* | *p<0.1; **p<0.05; ***p<0.01 |

### 2. Overall Mortality vs. Insolation

The dataset shows basically no correlation between the mortality rate (evaluated at the end of a month) and the overall insolation received by the country area in that entire month (Pearson’s r(35)=-0.132, P=0.435).

We plotted the regression line in Figure 2 for the completeness of the graphic. In the addition to the visually lack of a trend, the analysis of the residuals vs. leverage revealed some issues with the model: Slovakia is a high leverage point (with a high Cook’s distance); its exclusion from the analysis would significantly change the model. Quantile-quantile (Q-Q) plot analysis indicates that Slovakia, Italy and Belgium have more extreme values that would be expected from the rest of the data points. Therefore we conclude that in this dataset of 2020, in the European countries there was no apparent systematic correlation between insolation and COVID-19 mortality.

**Figure 2.**
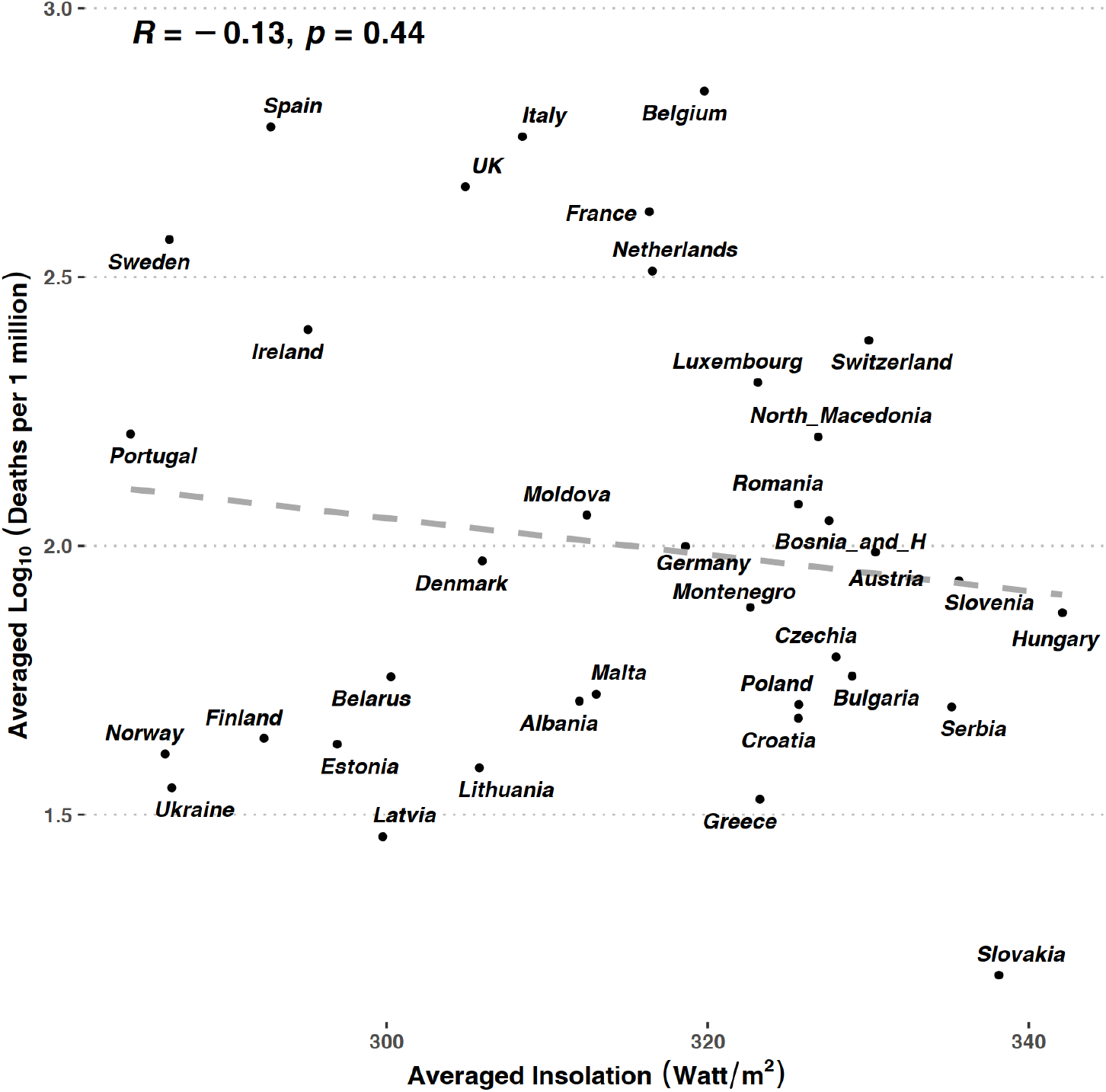
No correlation found between averaged mortality and averaged insolation (solar irradiance). The data points are the averaged values for all European countries included in the study during March – December 2020. The dashed line represent the linear model of the data (no fitting, see text); we included it as a guide for the eye.

As we noted in the Introduction, it is well known that the insolation (solar irradiance at the ground level) is primarily influenced by clouds and secondarily by other factors. It is expected that in general it should be a negative correlation between the cloudiness and insolation. We checked that this fact is indeed true in the analysed dataset: the insolation vs. cloud fraction has Pearson’s r(35)=-0.530, P<.001, and the adjusted R2=0.261 (averaged data for each country over the March-December interval, see Figure S7 in Supplementary material); in this averaged dataset, about 26 % variance of insolation is related to cloud fraction. The other confounding factors that influenced the variation are unknown, and we tried to address some of the possibilities in the Discussion section. A combined model of mortality vs. cloudiness and insolation (with or without interaction) did not significantly change the above results.

### 3. Monthly analysis

We also wanted to make sure that the above results were not the result of chronological influence over the chosen factors. As a preliminary check, we verified the independent variation in time of these variables; these are reported in Supplementary material, Figures S.4 – S.6. The overall mortality rate (Figure S.4) increases continuously. The cloudiness seem to be randomly distributed in the interval analysed (Figure S.5); this is expected because the seasonal temporal variability in the cloud cover is usually ∼30%, with bigger differences between extreme seasons (29). The average solar insolation in European countries month-by-month (Figure S.6) loosely follows the expected sinusoidal pattern due to seasonal changes (10).

After these checks we verified the correlations of these factors (cloud fraction, insolation) with mortality at monthly intervals (Figure 3).

**Figure 3.**
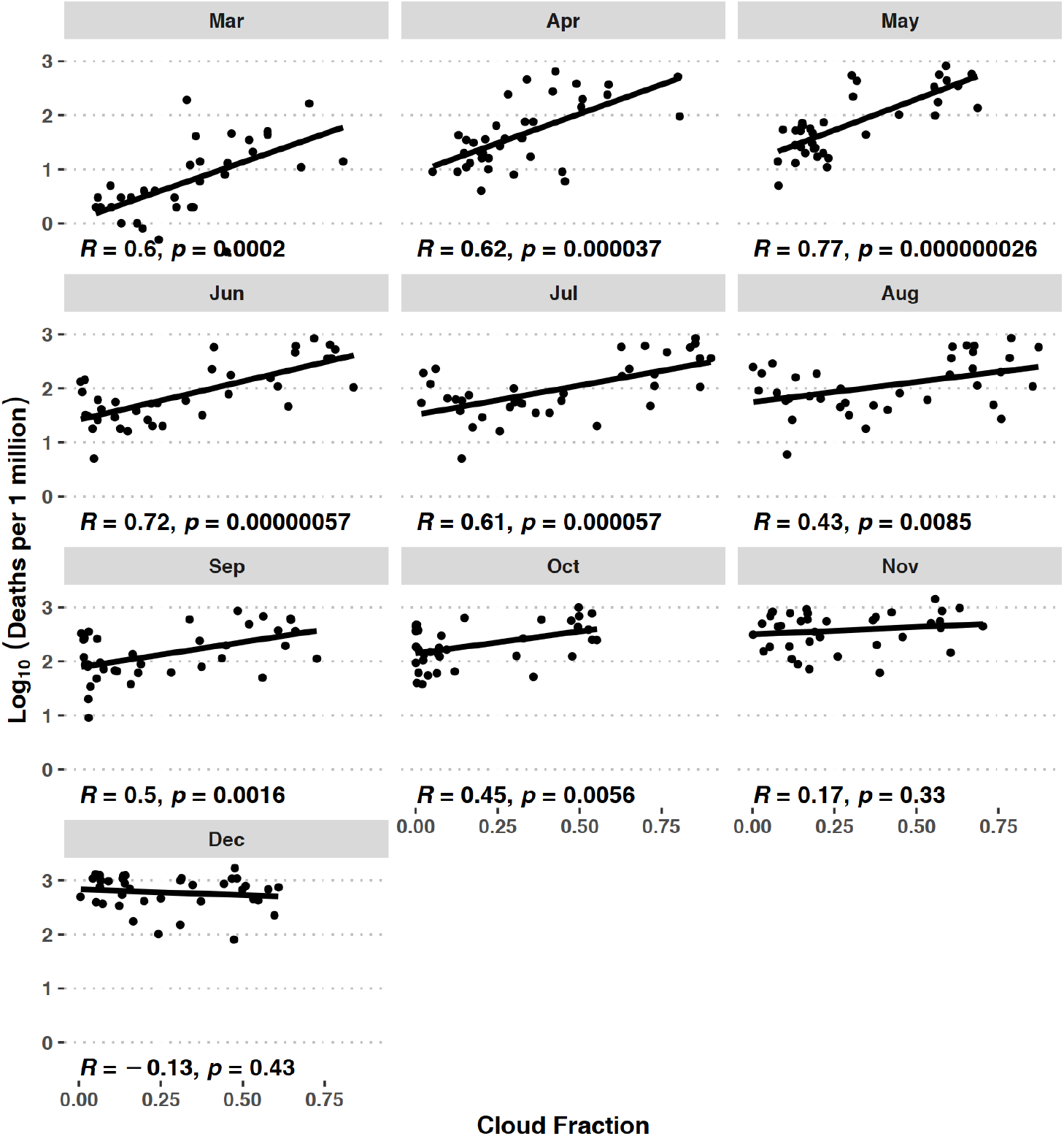
Monthly correlation between mortality and cloudiness. The data points are the values for all European countries included in the study for each month of the interval studied (March – December 2020). Cloudiness is expressed as the cloud fraction, which ranges between 0.0 (completely sunny sky) and 1.0 (clouds completely obscure the sun).

#### Mortality vs. Cloudiness over time

The dataset shows a positive and statistically significant correlation between the mortality rate (evaluated at the end of each month) and the overall cloudiness in that entire month (Pearson’s r listed in each monthly panel, Figure 3).

The linear models that fit the data shown in Figure 3 are presented in Table 2. In continental Europe, from the beginning of the epidemic, approximately 17% (October) to 59% (May) of the variance in COVID-19 mortality/million appears to be predicted by the cloudiness fraction of the sky in the March – October 2020 interval. The exception are November and December 2020 where there is no statistically significant trend.

**Table 2.**
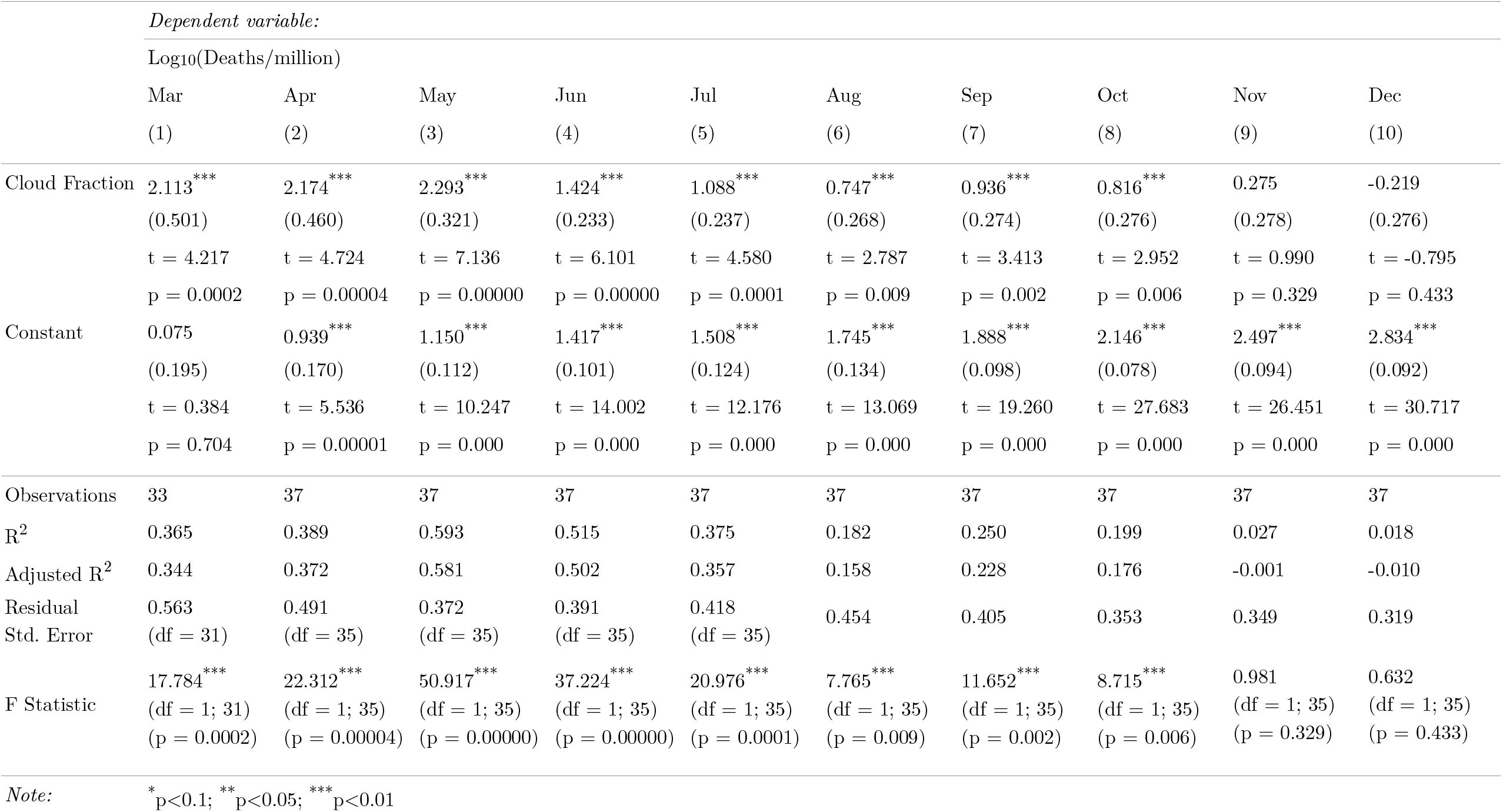
The statistics of the linear models of monthly mortality vs. cloudiness.

All the models for the months March – October passed the quality criteria listed in the Calculation section; the residuals of all the models appear to be homoscedastic, the Breusch-Pagan test yielding p-values>0.05 (range 0.121…0.955). All models passed a Durbin-Watson test for the autocorrelation of disturbances with a range of d-statistic=1.744…2.330, p-value>0.05, range 0.224…0.851).

The were no statistically significant correlation for November 2020 (P=0.33) and December 2020 (P=0.43). The mortality is similarly high in all countries; see Discussion for a possible explanation of this result.

#### Mortality vs. Insolation over time

For the dataset analysed we found no consistent correlation between the insolation and mortality in the month-by-month analysis. The Pearson’s r values were negative (with the exception of March) but the models did not pass the stringent criteria defined in the Calculation section.

### 4. Mortality vs. Latitude

The dataset (Figure 4) shows no correlation between the mortality rate and the latitude of the European countries for the entire interval studied (Pearson’s r(35)=-0.085, P=0.618). We used the “country centroid” (21) latitude as the single number that defined the latitude of a country (as discussed in the Methods section).

**Figure 4.**
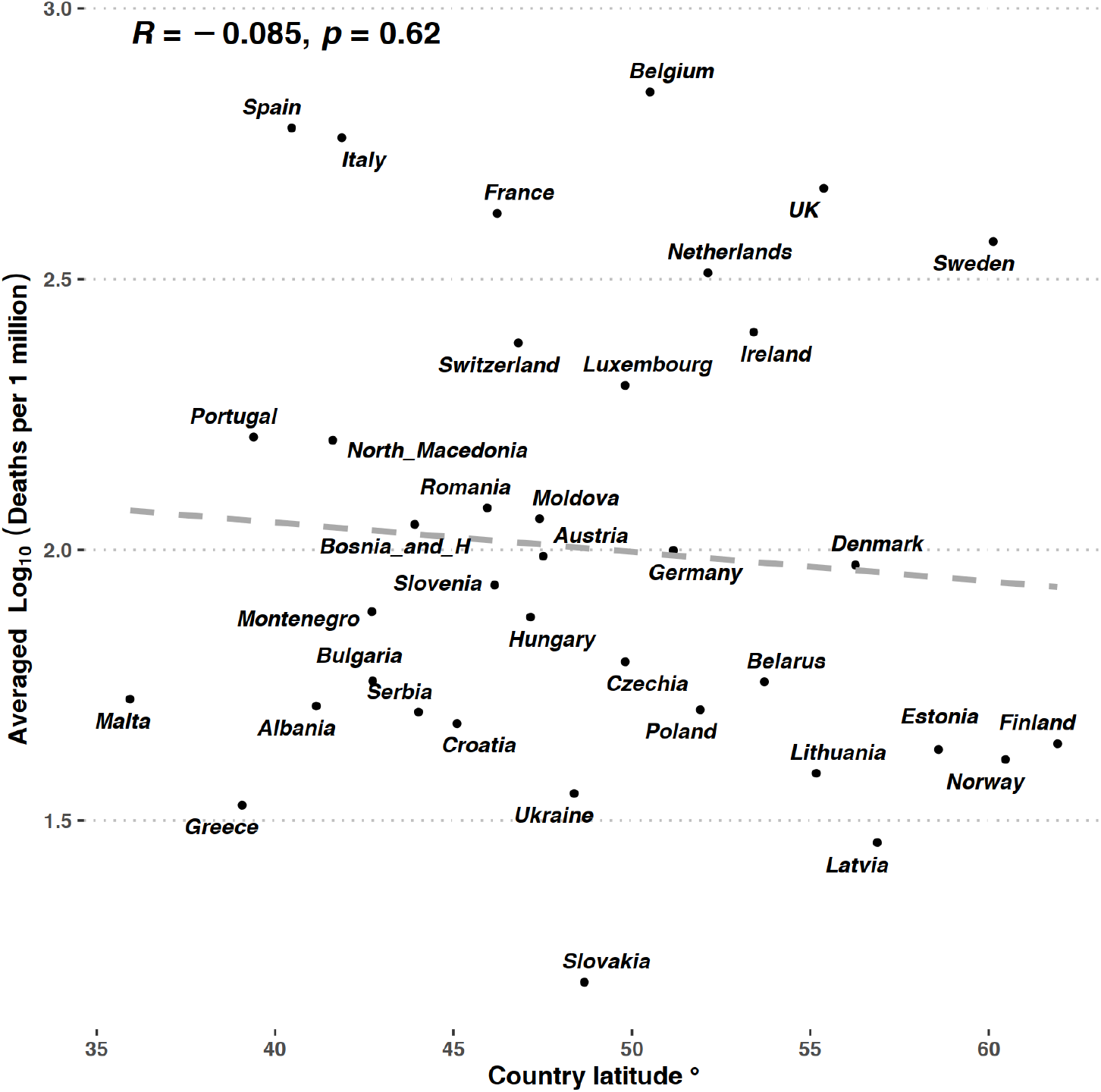
COVID-19 mortality and latitude of the European countries. No correlation found; the data points are the averaged mortality values for all European countries included in the study during March – December 2020. The dashed line represent the linear model of the data (no fitting, see text); we included it as a guide for the eye.

## Discussion

Our results show that in 2020 (the first year of the pandemic in Europe), the COVID-19 mortality was *not* related to the latitude or the amount of sunlight received; countries were affected regardless of their geographic position.

### Latitude

Our findings contradict some previously reported results about the existence of a latitude-mortality link (for example (2,5) and others). We think that our results is due to the fact that we used a rather more stringent definition of the “latitude of a country” (i.e. centroid definition, see Methds), rather than a broad geographical area (2) (broad geographical areas include large bodies of water, uninhabited) or the coordinates of the capitals (1) (capitals of the countries are not always the most populated cities).

### Insolation

For the whole interval (March – December 2020) we did not find a statistically significant correlation between the measured amount of sunlight and mortality. Previously published results supported the hypothesis that a greater amount of sunlight might have some beneficial effects (30). However the lack of month-by-month consistency and the low quality of the regression model suggests that the absolute amount of sunlight does not seem to have a significant impact on mortality.

Together, these two results seem to suggest that the mortality-vitamin D hypothesis (lower sunlight at higher latitudes leads to lower vitamin D levels, which leads to higher mortality) is perhaps incomplete. As a note of interest, other studies found that serum vitamin D correlates stronger with skin tone than with latitude (higher melanin content effectively prevents UV photons from initiating synthesis); see for example Åkeson et al. (31) for a study of vitamin D variation at a single latitude (Sweeden), Martin et al. (32) for a general meta-analysis, and a review of vitamin D and COVID-19 (33).

### Average Cloudiness

Unexpectedly, we found that among the factors studied (solar irradiance, latitude, cloud fraction), the cloudiness has the strongest positive correlation with COVID-19 mortality. This has been, to the best of our knowledge, unreported up to now.

Even if the average cloudiness correlation with mortality is small (Pearson’s r(35)=.68) is statistically significant. We suggest the following possible explanations for this unexpected finding of cloudiness influence on mortality:

#### 1) Heavy cloudiness is linked with colder outdoor surfaces, which might aid virus survival

A higher cloud cover quickly cools down the outdoor surfaces, especially at lower latitudes (34). Colder surfaces facilitate the survival of SARS-Cov-2 (35). As observed by Heneghan & Jefferson (36), the risk of incidence was higher in days with lower temperature (citing a study of a previous SARS epidemic by Lin et al, 2005 that observed an 18-fold increase in incidence in lower temperature days than in higher temperature days (37)); the plausible increase in actual incidence might thus lead to an increased number of deaths. As an easy-to-spot example for this unexpected finding, in the spring of 2020 Spain was cloudier than Norway (see Supplementary Material, Figures S.2 and S.1 for cloudiness and mortality charts).

#### 2) Reduced evaporation rate

We speculate that in addition to lowering temperature, a cloudy sky may drive down the evaporation rate (via two mechanisms: lower solar energy reaching the ground and perhaps by increasing the relative air humidity). This might favour the stabilization of the infectious droplets and enhance viral propagation (as suggested by Sajadi (2)). This seems to be supported by the evidence that coronaviruses survive better in colder environments but contradicts the finding that higher humidity impairs the transmission (3). This is highly speculative because in this dataset we could not check for actual humidity in the atmosphere (and to further complicate the interaction, we note that rainy clouds do increase the humidity).

#### 3) Moderate atmospheric pollution may be linked to both cloudiness and mortality

Even moderate pollution in the atmosphere helps clouds form (38), and it was also previously found that airborne pollution was linked to worse outcomes for the COVID-19 patients (see for example (39) for documented pollution effects in Italy; other similar studies are currently under review). Therefore, it might be that cloudiness data could be actually a proxy for pollution data (which is not available for all countries).

#### 4) Behavioural changes due to cloudiness

We suggest that an additional simple hypothesis is that overly cloudy weather (a higher cloud fraction) might change behavioural choices at large scale, nudging people to spend more time indoors rather than outdoors. It was suggested that spending more time in closed environments facilitates the secondary transmission of COVID-19, thus driving up the rate of infections (Nishiura et al., 2020, under review). This explanation seems to be consistent with the observation that living in overcrowded conditions is associated with poorer outcomes (8) and with the observation that some other environmental factors are at play in addition to the lack of sun-induced vitamin D synthesis (9).

The results show a difference in the relationships of COVID-19 deaths to cloud fraction versus solar radiation. This could perhaps be explained by a higher net amount of infrared radiation that reaches the ground surface when higher clouds are present compared with the presence of lower clouds (40,41). Our study could not distinguish between the types of clouds constituting the cloud fraction.

### Month-by-Month analysis of Cloudiness

We broke down the analysis of cloudiness vs. mortality in a month-by-month follow-up (Fig.3; Table 2). For 8 out 10 of the months studied, the positive correlation between the two remains significant and recurrent. This changed only in the last two months (November and December 2020) when the correlation was lost. We noticed that in these last two months there was an enormous amount of infections in all the countries (super-exponential spread). We think that in these last two months the mortality was driven up by the sheer amount of exposure of the population, and this dwarfed the rest of the confounding factors (including cloudiness).

### Limitations

We are aware that this study has limitations: we did not investigate the precipitation rate, wind velocity, air pressure, air pollution and density (these are factors that are under scrutiny for their possible impact on COVID-19 epidemiology). We acknowledge that our observational retrospective study is limited: temporal autocorrelation cannot be excluded over longer periods of time. The possibility of spatial autocorrelation was not researched by this study. Cloudiness might be a confounding factor in the previous studies that related vitamin D synthesis to latitude and sunlight. The authors support the current guidelines that in patients with vitamin D deficiency this should be treated irrespective of any link with respiratory infections. We are also aware that different countries took different governmental responses to the COVID-19 crisis, which has lead to different epidemiological outcomes. Last, we urge the reader not to extrapolate these results because we only investigated these variables in 37 European countries, not in the entire world. Additionally, this investigation was time-limited for the first year of the pandemic in Europe (2020), for 10 months. This period correspond to the first and second waves of pandemic infections, when most of the population was immunologically naive to the virus (unvaccinated, previously uninfected). We hope that these results will bring a warning about the possible impact of extended cloudiness on COVID-19 transmission.

## Conclusions

The data from the European continent in the spring-winter of 2020 suggest that the atmospheric cloudiness over a longer period (i.e., a month) seems to explain about a third of the variance of coronavirus (COVID-19) mortality; a higher degree of sky cloudiness in a month seems to be correlated with an increased mortality rate in that month. This knowledge might help advise public health policies related to COVID-19 mitigation and control; we suggest increased vigilance and increased frequency of sanitation of outdoor high-risk surfaces during cloudy weather.

## Data Availability

We made available the data analysed in this study at the Zenodo open-access repository

https://doi.org/10.5281/zenodo.4266757

## Acknowledgements

Author contribution: O.S. and A.I. jointly wrote the text, O.S. provided clinical background expertise, A.I. performed the formal analysis, and V.B. assisted with data curation and storage. We gratefully thank Prof. Constan?a Ganea, Dr. Octavian Călinescu and Dr. Marius-Cristian Frunză for careful reading of the manuscript and valuable insights and suggestions during the writing and revising of the manuscript.

## Data Availability Statements

The data that support the findings of this study are openly available (16,19–22). We made available the full data-set analysed in this study at the Zenodo open-access repository (https://doi.org/10.5281/zenodo.4266757).

