## Supplementary Material for "COVID-19 mortality: positive correlation with cloudiness but no correlation with sunlight and latitude in Europe"

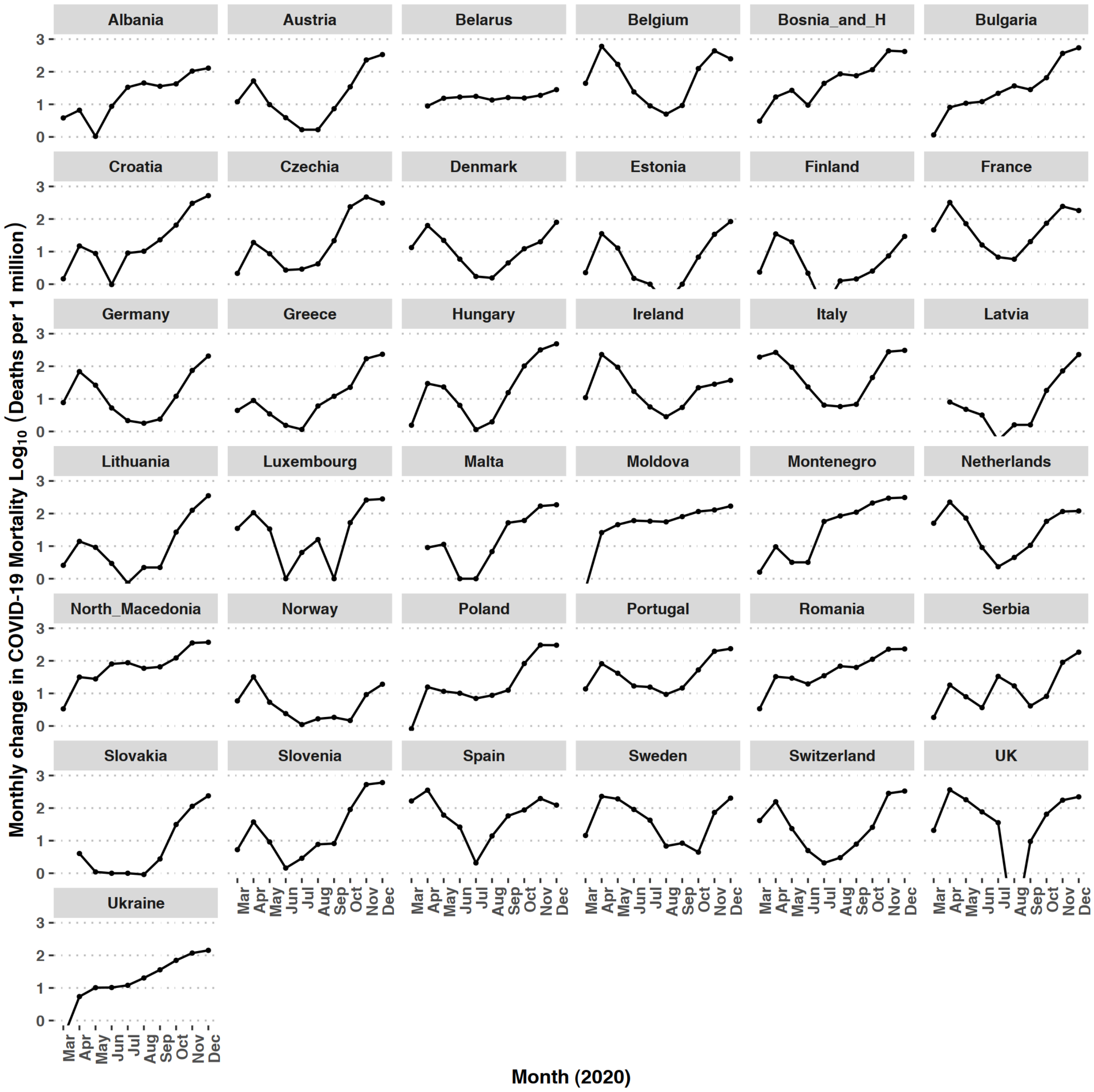


*Supplementary Figure S1.*
*COVID-19 deaths per 1 million population, in continental Europe, in 2020.*
*Each dot is the value of the deaths/million population at the end of each month. The lines are drawn to help guide the eye. Epidemiological data about COVID-19 deaths is available for most countries starting March 2020.*

A note for UK data: in August 2020 the health authorities UK changed the way the COVID-19 deaths were reported, and this shows as a visible gap. More information about this change can be found on an official statement of UK Health Security Agency, here: https://ukhsa.blog.gov.uk/2020/08/12/behind-the-headlines-counting-covid-19-deaths/

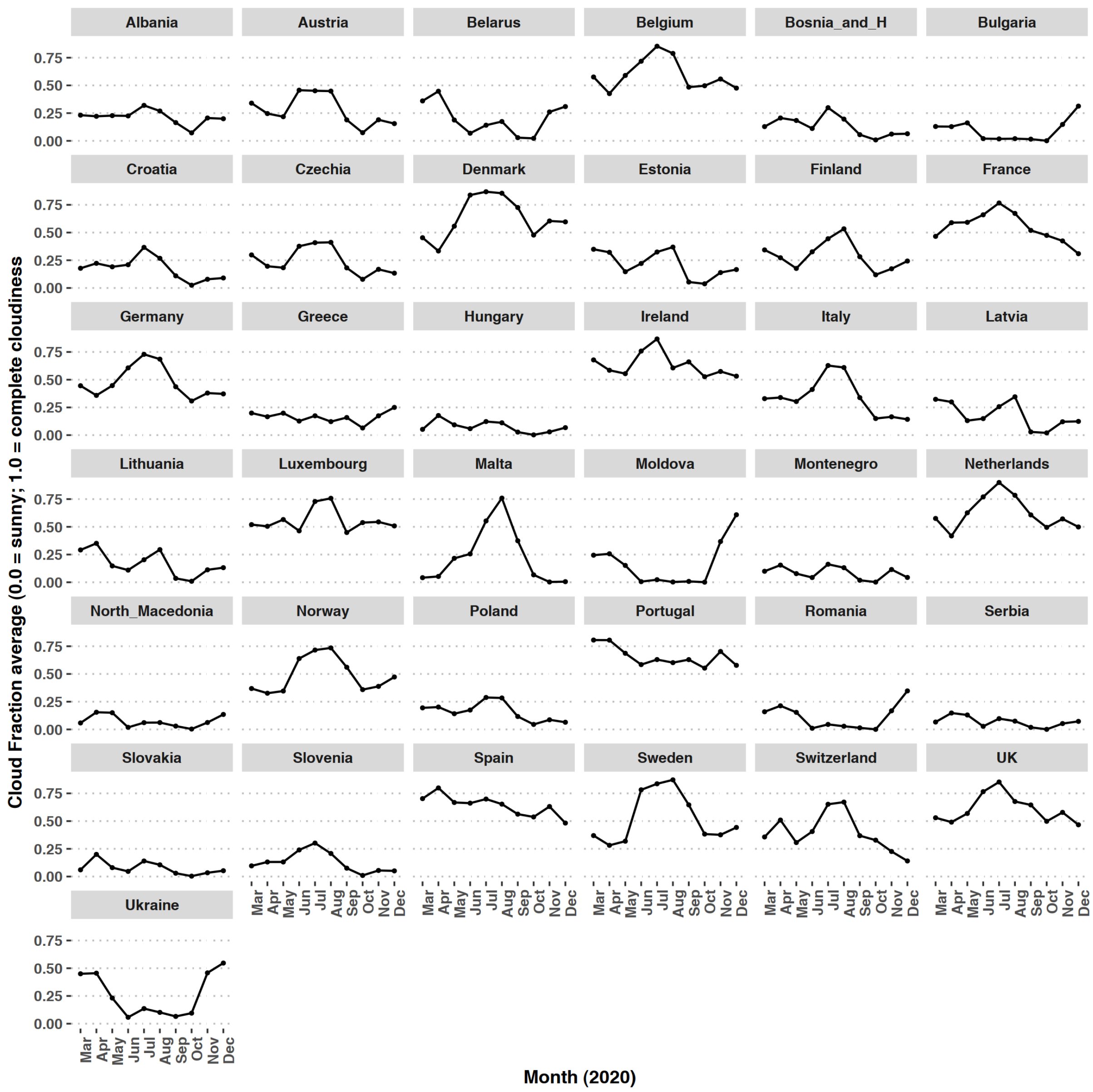


*Supplementary Figure S.2.*
*The measured cloudiness in each European country.*
*Each dot represents the average of cloud fraction for the geographical area of the country, during the respective month. A cloud fraction of 0.0 means a completely sunny sky; 1.0 means a completely clouded sky. The lines are drawn to help guide the eye.*

Authors' commentary: there is a huge variability of the cloudiness (as naturally expected); in 2020, some countries experienced a low cloudiness throughout the year (i.e. Greece), but others at similar latitudes had unexpectedly higher cloudiness (i.e. Spain, Portugal), etc. It is interesting to compare these charts with the insolation values (the next image).

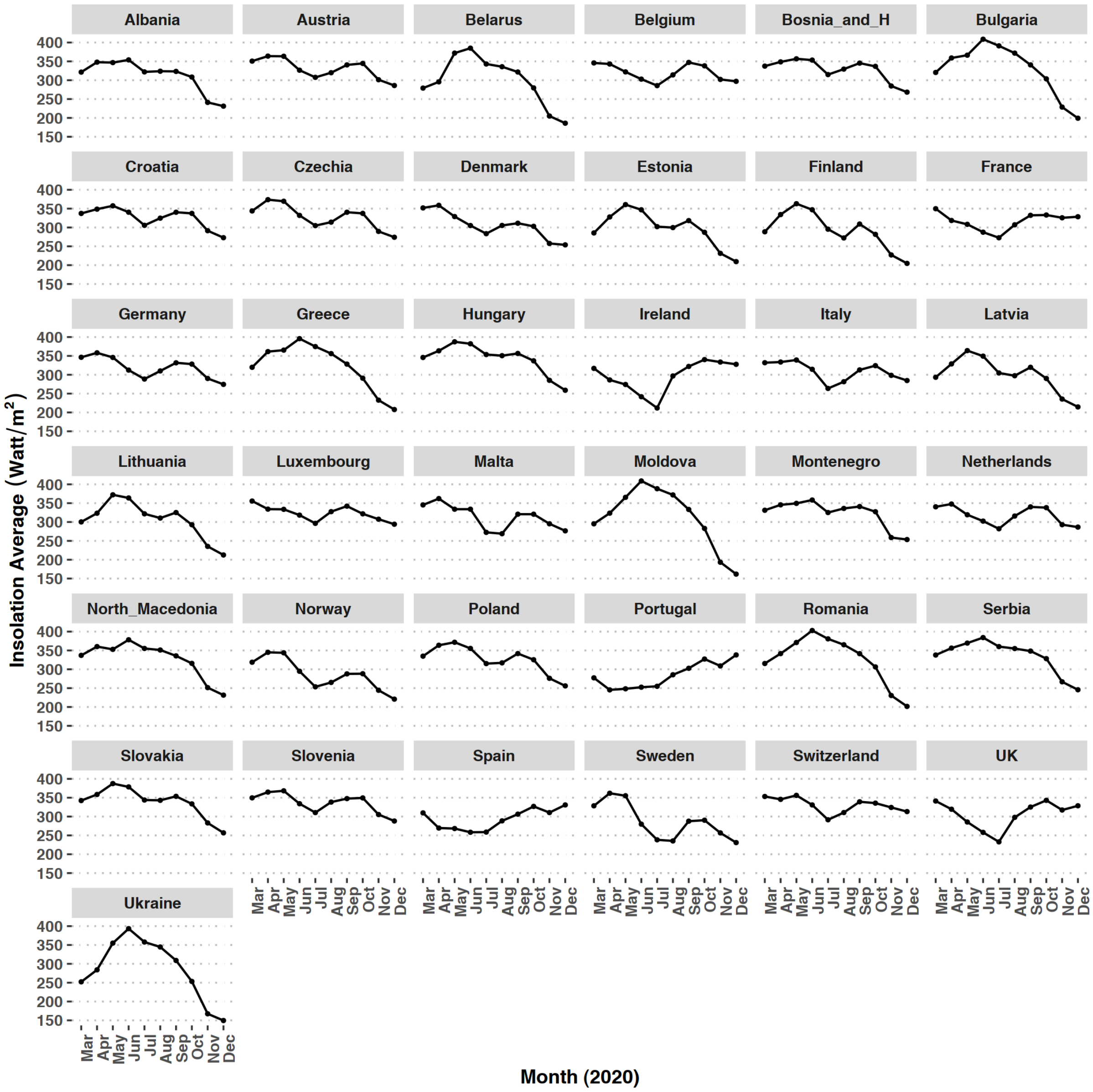


*Supplementary Figure S.3.*
*The average insolation in each country.*
*Each dot represents the averaged measured energy flux from the sun over the geographical area of the country, expressed in watt/$m^2$. The lines are drawn to help guide the eye.*

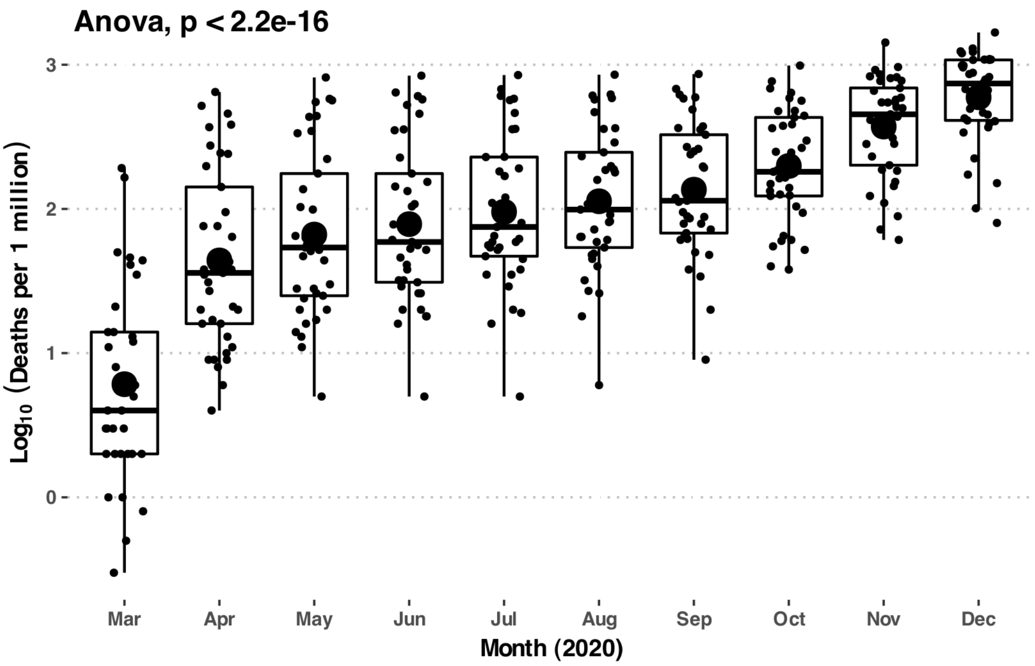


*Supplementary Figure S.4.*
*The boxplot of the average mortality in European countries in each month of the studied interval. The big central dots represent the means of each group; the smaller dots are the actual data points, automatically jittered for clarity.*

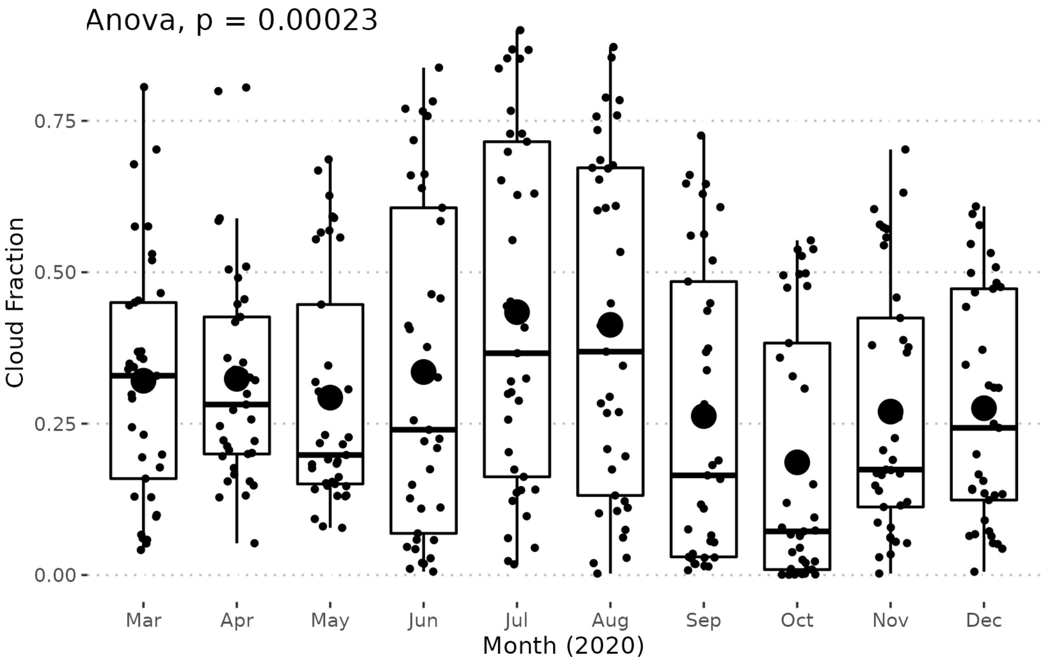


*Supplementary Figure S.5.*
*The boxplot of the average cloudiness in European countries in each month of the studied interval. The big central dots represent the means of each group; the smaller dots are the actual data points, automatically jittered for clarity.*

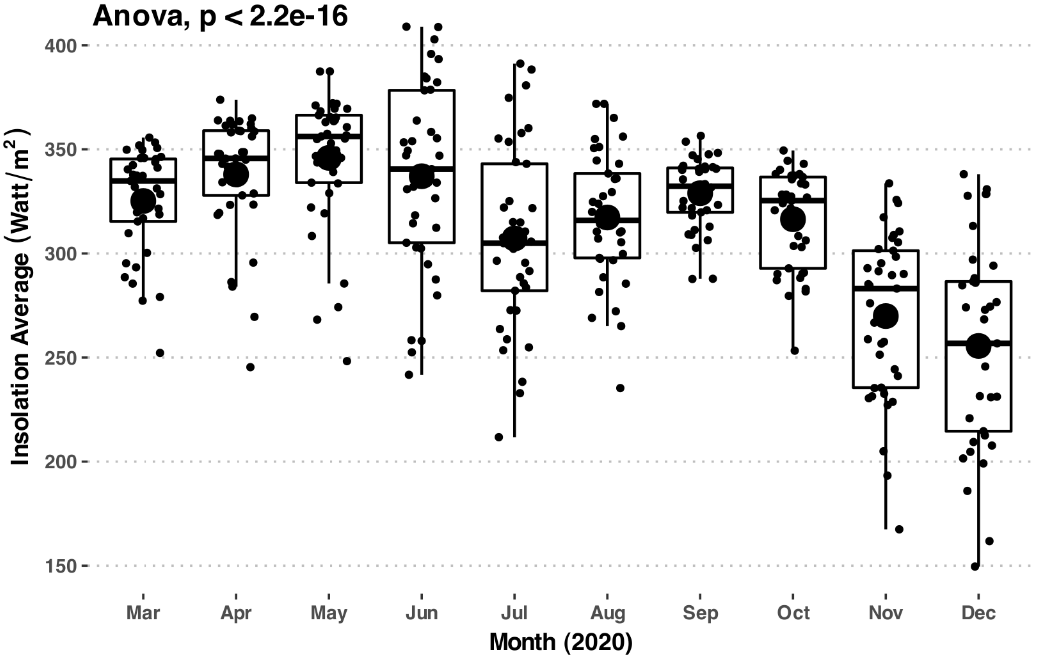


*Supplementary Figure S.6.*
*The boxplot of the average insolation received by European countries in each month of the studied interval. The big central dots represent the means of each group; the smaller dots are the actual data points, automatically jittered for clarity. There is a significant difference in insolation month-by-month (as expected due to seasonal change, see text).*

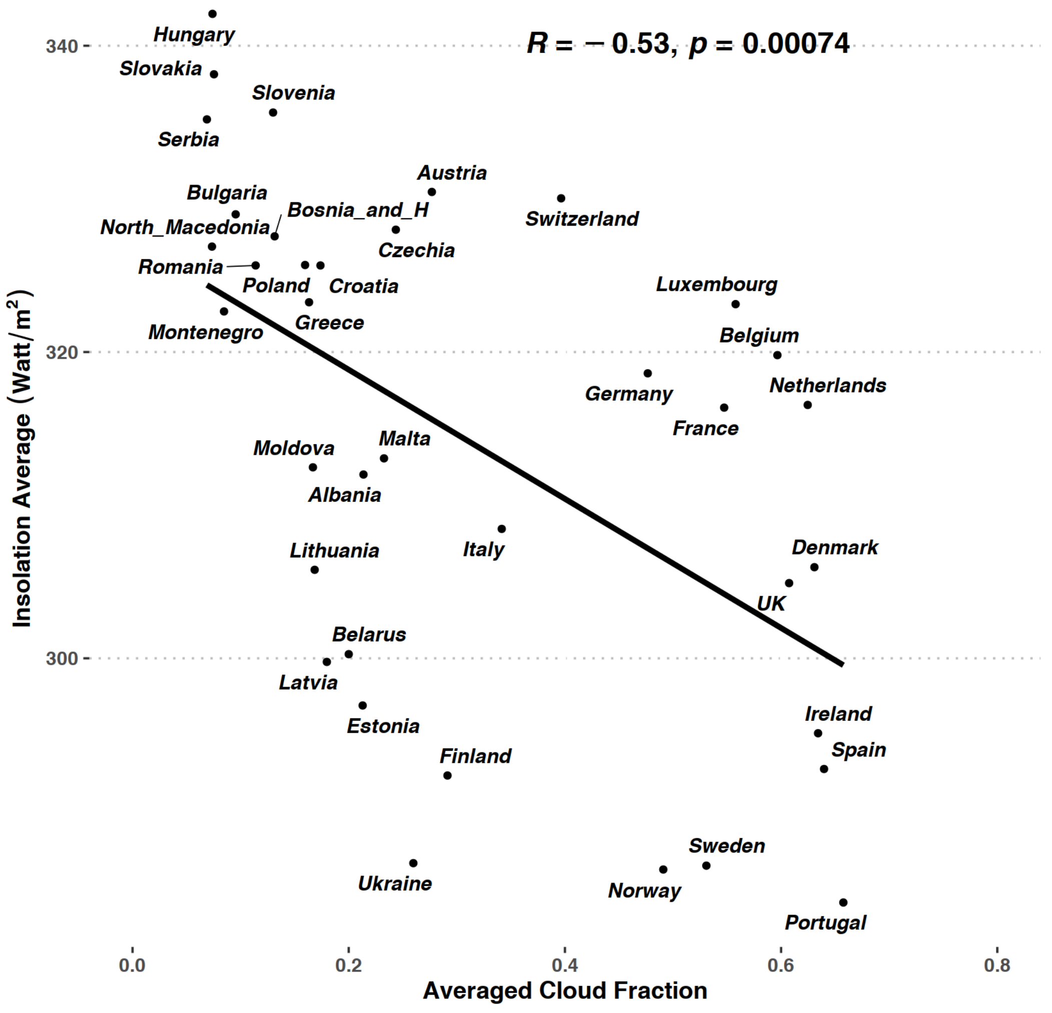


*Supplementary Figure S.7.*

*Per country Insolation vs. Cloud fraction (averaged values for the March – December 2020). As is expected, higher cloudiness correlates with lower insolation, but the correlation of the measured data has a lot of variance (there are many confounding factors that change the insolation at the ground level, in addition to cloud fraction). Pearson's r(35)=-0.530, P<.001, the adjusted $R^2$=0.261; the model passes the Breusch-Pagan test (p = 0.511). The statistics of the model are given in the following table.*

***Table S1. Supplimentary Table for figure S.7, with the linear model fitting the data***

| | *Dependent variable:* |
|---|---|
| | Insolation Average |
| Averaged Cloud Fraction | -42.164*** |
| | (11.399) |
| | t = -3.699 |
| | p = 0.001 |
| Constant | 327.277*** |
| | (4.241) |
| | t = 77.179 |
| | p = 0.000 |
| Observations | 37 |
| $R^2$ | 0.281 |
| Adjusted $R^2$ | 0.261 |
| Residual Std. Error | 14.110 (df = 35) |
| F Statistic | 13.683*** (df = 1; 35) (p = 0.001) |
| *Note:* | *p<0.1; **p<0.05; ***p<0.01 |
